# PABformer: Multi-Channel Transformer-Based Physical Activity Representation Learning from Wearable Accelerometry for Prediction of Parkinson’s Disease

**DOI:** 10.1101/2025.08.12.25333460

**Authors:** Kai He, Liuyi Fan, Hanyu Qian, Chunyu Li, Meng Wang

## Abstract

Parkinson’s disease (PD) is a prevalent neurodegenerative disorder whose early diagnosis and prediction remain challenging due to the delayed onset of overt motor symptoms and the complexity of long-term behavioral changes. Wearable accelerometers provide a scalable and non-invasive approach for monitoring physical activity (PA) in free-living environments and have emerged as a promising tool for PD prediction. However, existing accelerometer-based methods often rely on coarse summary statistics or models with limited ability to capture long-range temporal dependencies in multi-day accelerometer recordings. In this study, we propose PABformer, a multi-channel Transformer framework designed for long-term PA behavior representation learning from accelerometer data. PABformer introduces a channel-separation strategy to disentangle heterogeneous activity streams and leverages self-supervised pretraining to learn generalized behavioral representations. Using week-long accelerometer recordings from 96,463 participants in the UK Biobank, we pretrained PABformer and subsequently fine-tuned it for demographic attribute inference, PD diagnosis, and incident PD prediction. We compared PABformer against traditional statistical, machine learning approaches, recurrent deep learning models, and a standard Transformer without channel separation. Experimental results demonstrated that PABformer achieved superior performance in both PD classification and survival prediction tasks, as measured by precision–recall area under the curve (PRAUC) and concordance index (C-index), respectively. In addition, Grad-CAM analysis improved interpretability by identifying salient channel contributions associated with PD-related behavioral patterns. These findings demonstrate the potential of foundation models for wearable accelerometer data to advance early PD detection, disease risk prediction, and future precision health applications.

## I. Introduction

Parkinson’s disease (PD) is a prevalent neurodegenerative disorder that poses an increasing global health burden, particularly in aging populations. Its diagnosis remains complex and is largely based on clinical assessment of motor symptoms, which typically emerge only after substantial neuronal degeneration has occurred, leading to delayed and sometimes inaccurate diagnosis [1], [2]. Notably, many patients exhibit a prodromal phase characterized by subtle motor changes and non-motor symptoms years before clinical diagnosis, but these early signs are often difficult to detect reliably in routine practice [3]. These limitations underscore the urgent need for scalable, objective, and cost-effective approaches for early diagnosis and prediction.

Wearable accelerometers offer a promising solution by enabling continuous, non-invasive monitoring of movement patterns in real-world settings, thereby providing quantitative biomarkers for early detection. Accelerometry has emerged as a scalable, objective approach for assessing physical activity and motor behavior, transforming human movement analysis over the past decade and contributing to advances in both population health research and clinical care [4], [5]. These devices are widely adopted due to their accuracy, affordability, and accessibility [6], [7], [8], making them particularly well suited for large-scale and long-term monitoring [9], [10].

Current research on accelerometer-based approaches for PD diagnosis and prediction can be broadly categorized by the temporal granularity of physical activity (PA) signals. At short time scales (e.g., seconds to minutes), high-frequency accelerometry enables the extraction of detailed gait features—such as step variability, cadence, arm swing, and asymmetry—which have been shown to differentiate PD patients from healthy individuals and to predict disease conversion in the prodromal phase [11], [12]. These fine-grained motor signatures capture subtle abnormalities in movement dynamics that are not readily observable in clinical settings. In contrast, at longer time scales (e.g., days to weeks), accelerometer data are typically summarized into statistical descriptors of activity patterns, including the mean, standard deviation, and maximum of time spent in different PA categories such as moderate-to-vigorous physical activity (MVPA), light-intensity physical activity (LIPA), and sleep [13], [14], [15], [16], [17], [18]. Such aggregated features have demonstrated strong potential for identifying individuals at risk of prodromal PD years before clinical diagnosis [19]. However, these summary statistics may oversimplify complex behavioral patterns and temporal dynamics inherent in long-term activity data. This limitation highlights the need for more expressive and informative representations of long-term accelerometer data to fully exploit their potential for PD prediction. This work focuses on the representation learning of the long-term physical activity data.

To capture temporal dynamics in accelerometer data, functional principal component analysis (fPCA) has been widely used to represent activity trajectories as linear combinations of basis functions [20], [21]. While effective for summarizing overall trends, fPCA inherently emphasizes smooth variations and may obscure clinically relevant irregularities, such as abrupt movements (e.g., arm swinging) and sporadic physical activity patterns that are characteristic of PD. In contrast, deep learning approaches leverage nonlinear activation functions (e.g., ReLU and Tanh) to model complex thresholds and heterogeneous temporal patterns that linear methods may fail to capture [22]. Recurrent neural networks (RNNs) and long short-term memory (LSTM) networks have advanced time-series modeling [23], [24], but they remain susceptible to vanishing and exploding gradients, which limits their ability to capture long-range temporal dependencies [25]. These limitations are particularly problematic for continuous, multi-day accelerometer recordings, where clinically meaningful behavioral patterns may emerge intermittently across multiple temporal scales. Consequently, existing computational approaches provide only a partial characterization of the complex temporal structures embedded in long-term physical activity data.

Self-supervised learning (SSL) offers a powerful and scalable approach for extracting PA behavior representations from long-term accelerometer data. Unlike supervised methods that rely on large, labeled datasets, SSL leverages the inherent temporal structure of unlabeled time-series signals to learn generalizable and context-aware representations [26], [27]. Through pretext tasks—such as predicting future segments, contrasting temporally adjacent sequences, or reconstructing masked signal regions— SSL enables models to capture complex temporal dynamics, contextual variability, and personalized behavioral patterns without the need for manual annotation [28], [29], [30], [31], [32]. Transformers have emerged as a particularly effective architecture for implementing SSL in sequential data domains [33], [34], [35]. Their self-attention mechanisms are well-suited to learning from SSL tasks that require modeling long-range dependencies and global context [36], [37]. This capacity is especially advantageous for long-term accelerometer data, in which relevant behavioral patterns may extend over hours to days and require robust characterization of long-range interactions. Despite their success in general time-series applications (e.g., meteorology, traffic forecasting), analyzing long-term accelerometer data remains challenging, owing to their high sampling frequency and the heterogeneity of predominantly unlabeled activity patterns. While a recent study implemented a basic Transformer model with a patching technique on the NHANES accelerometer data (approximately 10,000 participants) for mental health prediction [38], it did not comprehensively address the issues of behavioral heterogeneity in long-range temporal modeling.

To overcome current modeling limitations, we propose PABformer, a multi-channel transformer framework tailored for PA behavior representation learning. PABformer is designed as a foundational model that explicitly disentangles heterogeneous activity streams, enabling more expressive and structured representations of long-term accelerometer data. The contributions of our work are summarized as follows:

- We introduce PABformer, a multi-channel transformer architecture that separates heterogeneous activity channels to better capture complex, long-term behavioral patterns in accelerometer data.
- We leverage the UK Biobank, utilizing high-resolution, week-long recordings from 81,463 participants to pretrain a foundational model for PA analysis.
- We fine-tune PABformer for downstream tasks including demographic attribute inference as well as PD diagnosis and prediction.
- We benchmark PABformer against a Transformer without channel separation, multiple traditional machine learning and deep learning models, demonstrating improved performance in PD diagnosis (PR AUC) and prediction using survival analysis (C-index).
- We enhance model interpretability by incorporating Grad-CAM to identify salient channel contributions, improving transparency and providing clinically meaningful insights.

The rest of the paper is organized as follows: Section II describes the study cohort and data for the analysis. Section III presents the proposed PABformer framework and implementation. Section IV reports the experimental results for demographic attribute inference as well as PD classification and prediction. Sections V and VI discuss the findings and study limitations, respectively. Finally, Section VII concludes the paper.

## II. Data Collection

### A. Study population

The UK Biobank is a population-based cohort that holds in-depth information on ∼500,000 participants aged between 40 and 69 years when recruited between 2006 and 2010 [39]. The cohort provides extensive multimodal data, including demographic characteristics, accelerometry measurements, and linked electronic health records.

This research is conducted under the approved application code 197947, with the data released to the University of Michigan. The informed consents of all the participants were obtained by UK Biobank.

### B. Datasets

#### 1) Accelerometer data

A total of 103,712 participants enrolled in the accelerometry sub-study of the UK Biobank. Participants were instructed to wear an Axivity AX3 wrist-worn triaxial accelerometer continuously for seven consecutive days between 2013 and 2015. Following the UK Biobank processing protocol [8], the raw 100 Hz triaxial signals underwent calibration, removal of gravitational and noise components, and detection of non-wear periods. The processed signals were subsequently converted into Euclidean Norm Minus One (ENMO) measures aggregated into 5-second epochs, with missing segments imputed by the UK Biobank pipeline. In addition, individual-level PA intensity measures were derived from the epoch-level accelerometry data. In this study, traditional PA features included derived accelerometry variables [40] (Category 1020), capturing hourly and daily proportions of time spent in sleep, sedentary, light-intensity, and moderate-to-vigorous physical activity, as well as acceleration summary variables (Category 1009), representing hourly and daily average PA intensities. The compared machine learning methods are based on these PA features. After quality control, 7,151 participants with poor calibration or insufficient wear time were excluded, resulting in 96,463 participants with reliable accelerometry data for analysis. The cohort was randomly divided into training (n = 81,463), validation (n = 5,000), and test (n = 10,000) sets. Unless otherwise specified, model pretraining was performed using the training cohort, while the validation and test cohorts were reserved for model selection and performance evaluation.

#### 2) Covariates

During the baseline assessment, the UK Biobank collected extensive demographic, lifestyle, and phenotypic information for all participants[39]. In this study, age at accelerometry assessment, sex and body mass index (BMI) were included as baseline covariates. Additional lifestyle factors considered included alcohol consumption frequency (daily or almost daily, three to four times per week, once or twice per week, one to three times per month, special occasions only, or never), smoking status (never, former, or current), and the number of days per week engaged in moderate physical activity (0–2, 3–4, or 5–6 days per week).

#### 3) Health outcome data

Disease diagnosis information was obtained from the first occurrence records (Category 1712), while mortality data were extracted from the death registry fields 40001 and 40002 in the UK Biobank. According to the UK Biobank protocol, first occurrence records integrate information from primary care data, hospital inpatient records, death registry data, and self-reported medical conditions to determine the earliest documented diagnosis date.

For the classification analysis, following prior work [19], participants diagnosed with PD before or within two years after accelerometer assessment were classified as diagnosed PD cases. Participants who received a PD diagnosis more than two years after accelerometer collection were defined as prodromal PD cases. Individuals without any diagnosed nervous system disorder were included as controls.

For the survival analysis, time-to-event was calculated from the date of accelerometer assessment. Among PD cases, the event time was defined as the interval to first PD diagnosis, measured in months. For participants without PD who remained alive, follow-up time was calculated from accelerometer assessment to the censoring date. Death was treated as a censoring event; for participants who died without a PD diagnosis, follow-up time was defined as the interval to death. Participants with prevalent PD at the time of accelerometer collection were excluded from the survival analysis.

## III. PABformer Model

### A. Overview of PABformer

We developed PABformer, a multi-channel transformer framework designed to learn rich behavioral representations from accelerometer data and support diverse downstream tasks (Fig. 1). The PABformer model builds upon a time-series Transformer architecture composed of stacked Transformer blocks [29]. The model inputs consist of 5-second epoch ENMO measurements derived from wrist-worn accelerometers collected continuously in free-living environments over 7 consecutive days. For each participant, one week of continuous monitoring yielded 120,960 epoch-level observations, providing high-resolution longitudinal PA data for downstream modeling.

**Fig. 1.**
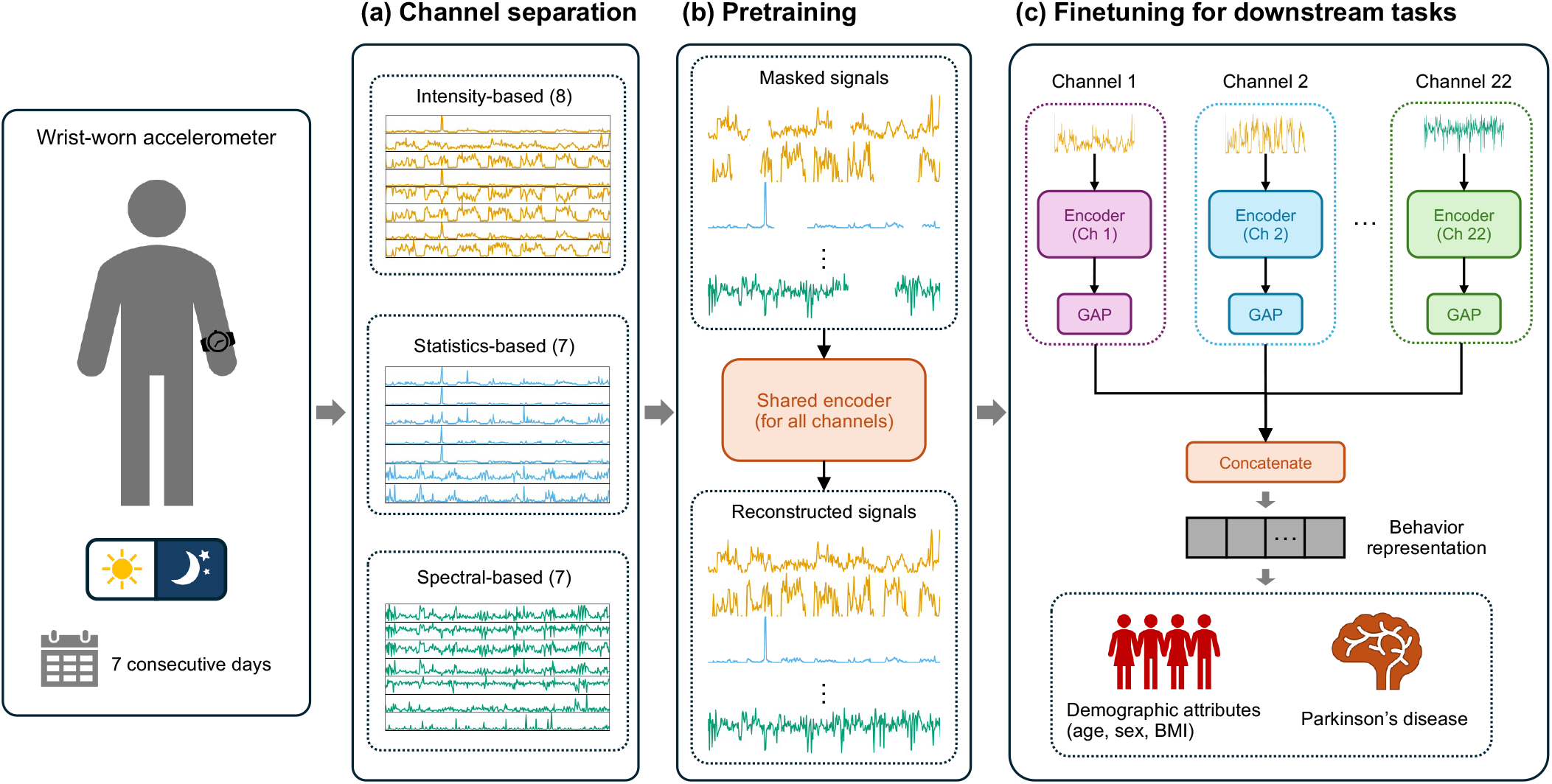
Overview of the Physical Activity Behavior Transformer (PABformer). Raw acceleration data were continuously recorded via a wrist-worn accelerometer over seven consecutive days in a free-living environment. (a) Channel separation of wearable sensor data. Raw acceleration data were processed into 5-second epochs to estimate physical activity intensity. For each individual, a total of 22 channels is derived from intensity data and categorized into intensity-based channels (8), statistics-based channels (7), and spectral-based channels (7). (b) Pretraining diagram of PABformer using multi-channel physical activity data. The core component of PABformer is a time series transformer, pretrained on PA data of 22 channels using a masked-patch prediction objective. (c) Finetuning diagram of PABformer for downstream tasks. A task-specific transformer encoder is adopted to extract relevant patterns. Embeddings from all channels are concatenated to form comprehensive behavior representations, enabling PABformer to learn task-specific representations.

The PABformer framework consists of three components: channel separation, pretraining, and finetuning for downstream tasks. In the channel separation (Fig. 1a), 22 channels are extracted from the ENMO signals to explicitly disentangle heterogeneous PA signals. This design allows each channel to be processed independently, enabling the model to capture nuanced, channel-specific patterns in parallel and better represent complex, long-term behaviors. During the pretraining stage (Fig. 1b), a shared Transformer encoder is applied across all PA channels. Each channel is independently masked and reconstructed using a self-supervised objective, allowing the model to learn generalizable representations from large-scale unlabeled accelerometer data. During the fine-tuning stage (Fig. 1c), the pretrained encoder is applied separately to each channel, and the resulting representations are concatenated to form a comprehensive behavioral embedding for each individual. These embeddings are then passed to task-specific output layers for downstream applications, enabling the model to learn channel-specific importance and task-relevant patterns.

### B. Channel separation

In the step of channel construction (Fig.1b), 22 PA channels were derived from processed 5s epoch PA signals for each individual. These channels fall into three categories: intensity-based channels, statistics-based channels, and spectral-based channels. For the intensity-based channels, the 5-second epoch data were categorized into sedentary behavior (SB; <30 mg), light-intensity physical activity (LIPA; 30-100 mg), and moderate-to-vigorous physical activity (MVPA; >100 mg) based on ENMO values. Total activity feature was defined as the sum of all ENMO values within a given time interval. The total SB, total LIPA, and total MVPA features were calculated by summing the ENMO values corresponding to each category. The SB count, LIPA count, and MVPA count features were computed by counting the number of epochs classified into each activity category. The transition count feature was calculated by counting the number of transitions between LIPA and MVPA. For the statistics-based channels, the standard variation, median, maximum, first-quantile, third-quantile, skewness, and kurtosis features were calculated to characterize the distribution of values within each time interval. For the spectral-based channels, the 5-second epoch PA signals were transformed into the frequency domain with the Fourier transformation. Subsequently, the following features were extracted: spectral centroid, spectral bandwidth, spectral entropy, spectral flatness, number of peaks, fundamental frequency, and dominant frequency. All the channels are extracted at 30-minute intervals, resulting in 336 timestamps to represent a full week of PA. For each individual, PA data were represented by 22 different channels, each consisting of 336 values. Each PA channel was normalized based on the population-level z-score. Non-wearing periods were imputed with zeros before feeding into transformer encoders.

### C. Self-supervised pretraining

In the pretraining stage, PABformer models were trained from all 22 channels to encode the PA data for each channel. For each channel, the PA data were divided into a patch sequence and masked randomly. The masked patch sequence was then processed by a shared time series transformer encoder, which generated the embeddings for each patch. A linear output layer was applied to predict the PA signals from the encoded patch representations. Only the masked patches and corresponding ground-truth signals were used to compute the masked-patch loss.

#### Setup

Let *X* ∈ ℝ^*N×C×T*^ denote the PA channels dataset constructed from accelerometer data, where *N* is the number of individuals, *C* = 22 is the number of channels per individual, and *T* = 336 is the length of the time series. The time series data of the *i*^*th*^ individual for individual channel *j*^*th*^ is denoted as *X*_*ij*_ ∈ ℝ^*T*^.

#### Patching

Each channel *X*_*ij*_ is partitioned into a sequence of non-overlapping patches of length *P*. The number of patches is given by 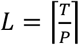, which is the result of dividing the sequence length *T* by the patch length *P*, rounded up to the nearest integer, to ensure complete coverage of the sequence. The *k*^*th*^ patch of *X*_*ij*_ is denoted as 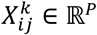. For each channel, the patch sequence was randomly masked at a fixed ratio, with the selected patches replaced by zero values. The masked patch is denoted by 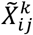 . The patching operation reduces the computation and memory usage of the transformer significantly, thereby enabling efficient modeling of long-term sequence data [29].

#### Embedding layers

Each patch is embedded via a learnable embedding function, and a learnable position encoding is added on:

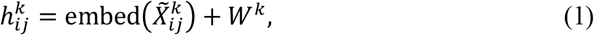

where *W*^*k*^ is the positional embedding for *k*^*th*^ patch. The embedded patches are then concatenated to a full patch sequence for the channel *j* of the individual *i*:

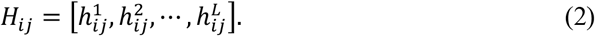

#### Transformer encoder

The patch sequence from each channel is passed through a shared time series transformer encoder to produce the channel representations individually:

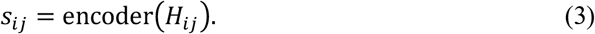

The transformer encoder contains multiple transformer blocks, each incorporating a multi-head self-attention mechanism to process the input patches [36], [41]. Within each attention head of *l*^*th*^ transformer block, the self-attention operation transforms the patch input 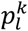 into query *Q*, key *K*, and value *V* using learnable projection matrices:

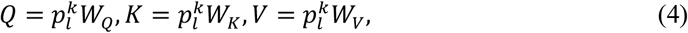

where *W*_*Q*_, *W*_*K*_, and *W*_*V*_ are the weight matrices for this head. The attention output is then computed as:

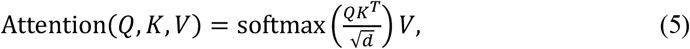

where *d* is the dimensionality of the query and key vectors, and the softmax operation ensures the attention weights sum to one. Finally, a linear projection head is used to reconstruct the original patches from the channel representations:

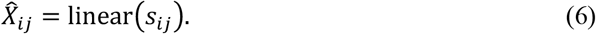

#### Loss function

The PABformer model is pre-trained using the mask-patch prediction loss [29]. A binary mask 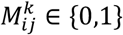 is used to indicate if *k*^*th*^ patch in *X*_*ij*_ is masked. The pretraining loss is defined by:

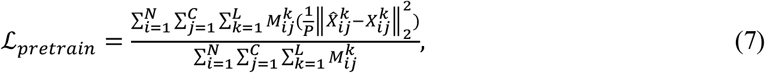

where *P* denotes the patch length, 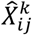 is the reconstructed patch, and 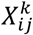 is the ground-truth patch.

### D. Downstream prediction framework

In the fine-tuning stage, PABformer is adapted to specific downstream tasks by leveraging channel-specific representations. The shared pretrained encoder is replicated and assigned to each PA channel, allowing each channel to be processed independently without masking. For each channel, the pretrained transformer encoder generates patch-level embeddings, which are then aggregated using global average pooling (GAP) to produce a compact channel-wise representation. These representations from all 22 channels are concatenated to form a comprehensive behavioral embedding for each individual. The combined embedding is subsequently passed through a task-specific output layer to predict the target outcome. By initializing each channel-specific encoder with shared pretrained weights, the model captures both generalizable patterns and channel-specific variations, enabling effective learning across diverse downstream tasks.

#### Channel-specific encoder

In contrast to the pretraining stage, each patch in this phase is embedded using a channel-specific embedding function and added with channel-specific positional encodings:

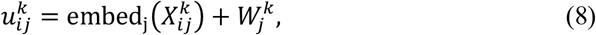

where 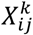 is the *k*^*th*^ patch input of *X*_*ij*_, and embed_*j*_ is the channel-specific embedding layer. After that, the embedded patch sequence 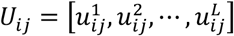 is passed through a channel-specific Transformer encoder:

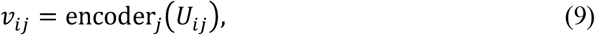

where encoder_*j*_ is the channel-specific transformer encoder for *j*^*th*^ channel.

#### Behavior representation

GAP operation is used to aggregate the embedding sequence into channel representations:

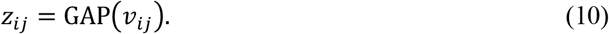

The PA representations across all channels are then aggregated to construct the behavior representation for individual *i*:

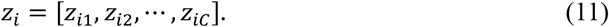

### E. Downstream tasks and evaluation

The behavior representations are transformed into the downstream outputs by a linear layer, with or without covariates, as follows:

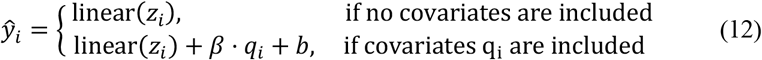

where ŷ_*i*_ denotes the prediction of individual *i* for a given downstream task, *q*_*i*_ is the covariate vector, *β* and *b* are weights and bias for covariates.

We evaluated PABformer on demographic attribute inference, PD classification and prediction.

For demographic attribute inference, PABformer models were finetuned to predict age, sex, and BMI without incorporating additional covariates. Age and BMI prediction were formulated as regression tasks optimized using mean squared error (MSE) loss and evaluated using mean absolute error (MAE) and coefficient of determination (*R*^*2*^). Sex prediction was formulated as a binary classification task optimized using cross-entropy loss and evaluated using accuracy and F1 score.

For PD classification tasks, we evaluated PABformer on three binary classification tasks: diagnosis, prodromal prediction, and screening (defined in Section II). PABformer models were finetuned using binary cross-entropy loss. Because PD classification tasks are highly imbalanced, performance was primarily evaluated using the precision–recall area under the curve (PR AUC) instead of the receiver operating characteristic AUC (ROC AUC) [42].

Incident PD prediction was evaluated using survival analysis incorporating accelerometer-derived representations and traditional covariates. A baseline Cox proportional hazards (Cox PH) model was first used to estimate covariate effects, with these coefficients fixed during the fine-tuning stage. PABformer was then fine-tuned using the negative partial log-likelihood loss function [43]:

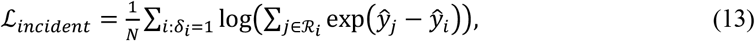

where *N* denotes the number of observed events in the batch, δ_*i*_ ∈ {0, 1} indicates whether an event occurred for individual *i*, and *R*_*i*_ is the risk set consisting of individuals still at risk at the time of the event *i, ŷ*_*i*_ denotes the predicted log-risk score of individual *i* defined in (12). Model performance for incident PD prediction was evaluated using the concordance index (C-index), inverse probability of censoring weighted concordance index (IPCW C-index), and 5-year area under the receiver operating characteristic curve (AUROC). The C-index measures the model’s ability to correctly rank survival times, with higher values indicating stronger concordance between predicted risk scores and observed event times. The IPCW C-index further accounts for potential bias introduced by censoring, providing a more robust assessment of survival prediction performance in the presence of incomplete follow-up data. The 5-year AUROC evaluates the model’s discriminative ability to predict whether a PD event will occur within five years after accelerometer assessment.

### F. Implementation details

The accelerometer dataset was randomly divided into training, validation, and testing sets containing approximately 85%, 5%, and 10% of participants, respectively. Unless otherwise specified, all experiments were conducted using identical data splits to ensure fair comparison across models.

PABformer model consisted of 6 Transformer blocks. Each block contained 4 attention heads. During pretraining, random patch masking was applied during wearing periods with a masking ratio of 0.2 to improve robustness against missing or irregular activity patterns. Both model variants were pretrained using the AdamW optimizer [44] for 200,000 steps with a batch size of 32, an initial learning rate of (5 × 10^*−4*^), and a linear learning-rate scheduler.

For demographic attribute inference tasks, including age, sex, and BMI prediction, PABformer model was finetuned for 50,000 steps. The learning rate was initialized at (1 × 10^*−5*^) for age and sex prediction, and (1 × 10^*−6*^) for BMI prediction. For BMI prediction, valid BMI measurements within six months of accelerometer acquisition were available for 2,907 participants. Age and BMI prediction tasks were optimized using MSE loss, whereas sex prediction was optimized using cross-entropy loss. For PD classification tasks, PABformer model was finetuned using a learning rate of (1 × 10^*−6*^) for 30,000 steps. To mitigate severe class imbalance in PD classifications, a balanced sampling strategy was adopted during training to increase the representation of positive PD cases within each mini-batch. Model selection and early stopping were performed based on validation PR AUC.

For PD survival analysis, PABformer was finetuned using a time-to-event objective optimized with partial likelihood loss. The model was trained for 15,000 steps using the AdamW optimizer with a learning rate of (5 × 10^*−6*^) and a batch size of 64. To address event imbalance, each mini-batch consisted of a mixture of censored and event samples, with the sampling ratio selected empirically using the validation set. The Breslow estimator [45] was used to estimate the cumulative baseline hazard function, from which individual survival probabilities and risk scores were derived.

To interpret model predictions, we employed an aggregated GradCAM-based visualization approach [46]. To illustrate the results, GradCAM was applied to the PD diagnosis task by randomly selecting 100 individuals diagnosed with PD within two years after accelerometer acquisition. Positive neural activations were backpropagated to compute gradients of PA representations before the global average pooling layer, generating patch-wise channel importance scores. Importance values were aggregated across individuals using a maximum operation and subsequently normalized by the global maximum score across channels.

Traditional machine learning baselines were implemented using Scikit-learn (v1.5.2) [47] and XGBoost (v2.1.4) [48]. Survival analysis was conducted using Scikit-survival (v0.23.0) [49] and pycox [43]. The PABformer models were implemented using PyTorch (v2.5.0) [50] and Transformers (v4.45.2) [51]. Data preprocessing was performed using NumPy (v1.26.4) and Pandas (v2.2.3). Figures were generated using matplotlib (v3.9.2) and seaborn (v0.13.2).

## IV. Experimental Results

### A. Demographic attribute inference

We evaluated the performance of PABformer models in demographic attribute inference, a commonly used benchmark for assessing pretrained foundation models [52], [53]. The model was finetuned to infer demographic attributes, including age, sex, and BMI. Specifically, we appended a fully-connected layer that maps the extracted behavior representations to the target demographic outputs.

PABformer models were first evaluated on the age estimation task. The PABformer models achieved a *R*^*2*^ score of 0.52, indicating that predicted ages reasonably reflected the chronological age for most individuals (Fig. 2a). Next, the models were assessed on the sex classification task, with a confusion matrix shown in Fig. 2b. PABformer models achieved high recall (>0.8) for both male and female classes. Finally, the models were applied to estimate BMI, where predicted BMI values showed a moderate correlation with measured BMI values (Fig. 2c).

**Fig. 2.**
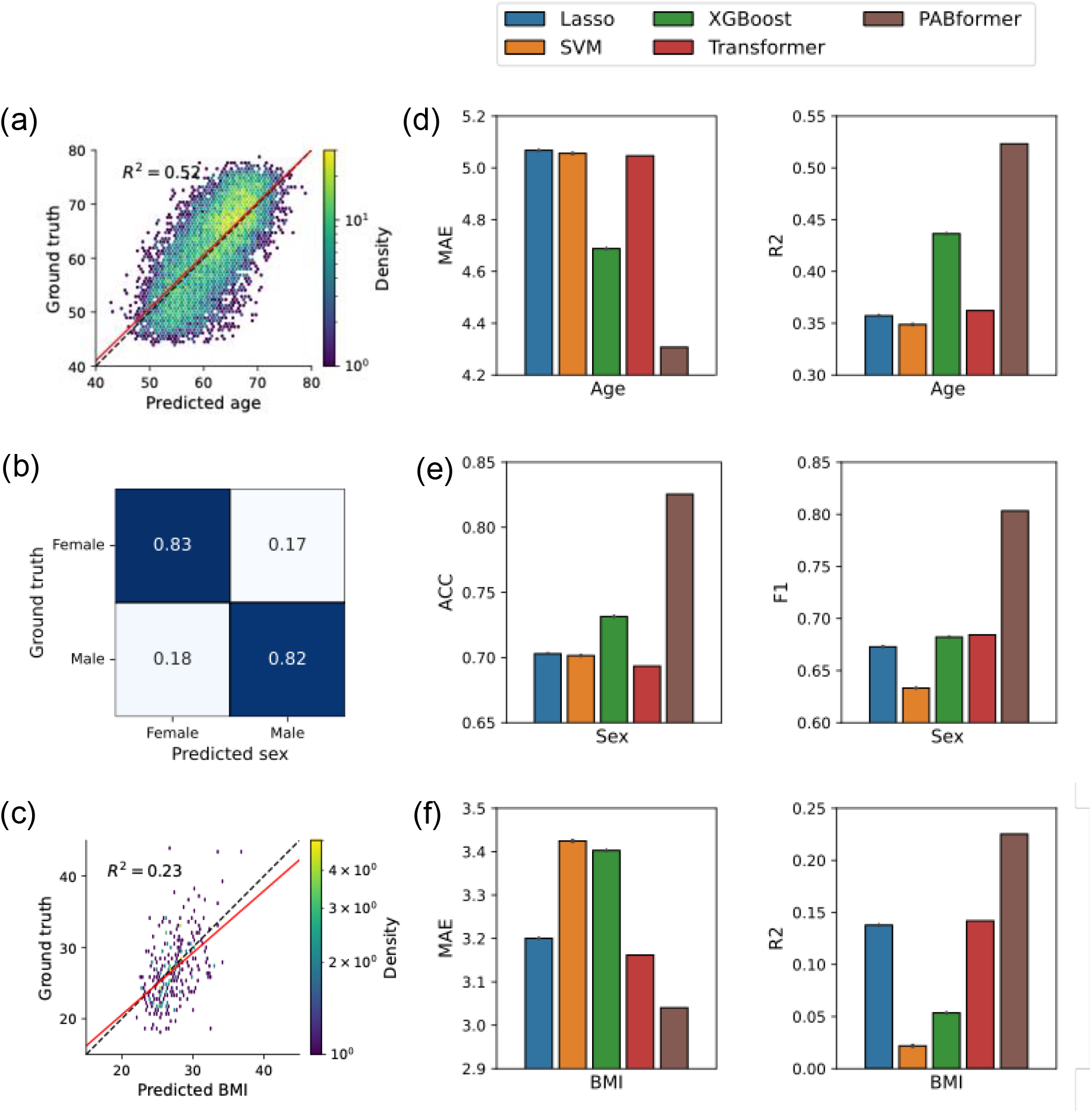
Demographic attribute inference using PABformer and model comparison. (a) Scatter plot illustrating the relationship between chronological age and age predicted by PABformer. (b) Confusion matrix between ground truth sex and predictions by PABformer. (c) Scatter plot illustrating the relationship between BMI and its prediction from PABformer. (d) Bar plots comparing the mean absolute error (MAE) and coefficient of determination (R2) for age estimation across various baseline models and PABformer. (e) Bar plots comparing the accuracy (ACC) and F1 for sex prediction across various baseline models and PABformer. (f) Bar plots comparing the MAE and R2 for BMI estimation across various baseline models and PABformer.

We compared the PABformer models with baseline models on the demographic attribute inference tasks (Fig. 2d-f. PABformer consistently outperformed all baseline models across all demographic inference tasks and evaluation metrics. Notably, PABformer achieved improvements of about 8 percentage points in *R*^*2*^ for age prediction, about 12 percentage points in F1 score for sex prediction, and about 8 percentage points in *R*^*2*^ for BMI prediction. In contrast, the Transformer model performed only slightly better than traditional machine learning baselines on the BMI estimation task and underperformed on the age and sex prediction tasks. These results highlight the effectiveness of the channel separation strategy employed in PABformer models.

### B. Model comparison on PD classification tasks

To comprehensively evaluate the effectiveness of PABformer for accelerometer-based PD classification tasks, we compared it against a diverse set of baseline approaches including the statistical method (fPCA), machine learning approaches (Lasso, SVM, and XGBoost), recurrent neural networks (RNN and LSTM), and a standard Transformer without channel separation [29][48].

For fPCA, each participant’s physical activity trajectory was represented using functional principal component scores derived from the 30-minute resolution accelerometer sequence. Lasso, SVM, and XGBoost were trained using 359 handcrafted physical activity features, including activity intensity, sedentary behavior, transition-related measures, statistical descriptors, and frequency-domain features. RNN, LSTM, and Transformer models were trained directly on the multichannel accelerometer-derived time-series inputs. The Transformer baseline employed the same input representation as PABformer but processed all channels jointly without channel separation.

All deep learning models were optimized using the AdamW optimizer with early stopping based on validation PR AUC. Weighted loss functions were applied during training to mitigate the severe class imbalance in PD-related prediction tasks.

The PR AUC results are summarized in Table I. PABformer consistently achieved the best overall performance across diagnosis, screening, and prodromal prediction tasks, reaching PR AUC values of 0.512, 0.429, and 0.214, respectively. The prodromal task is the most challenging, as all models show lower PR AUC compared with diagnosis and screening. Nevertheless, PABformer maintains the best performance in this setting, suggesting its ability to capture subtle pre-diagnostic behavioral changes associated with future PD risk. Notably, the largest relative gain over the Transformer baseline was observed in the prodromal prediction task, suggesting that channel separation becomes increasingly important when disease-related behavioral changes are weak and difficult to detect.

**TABLE I.**
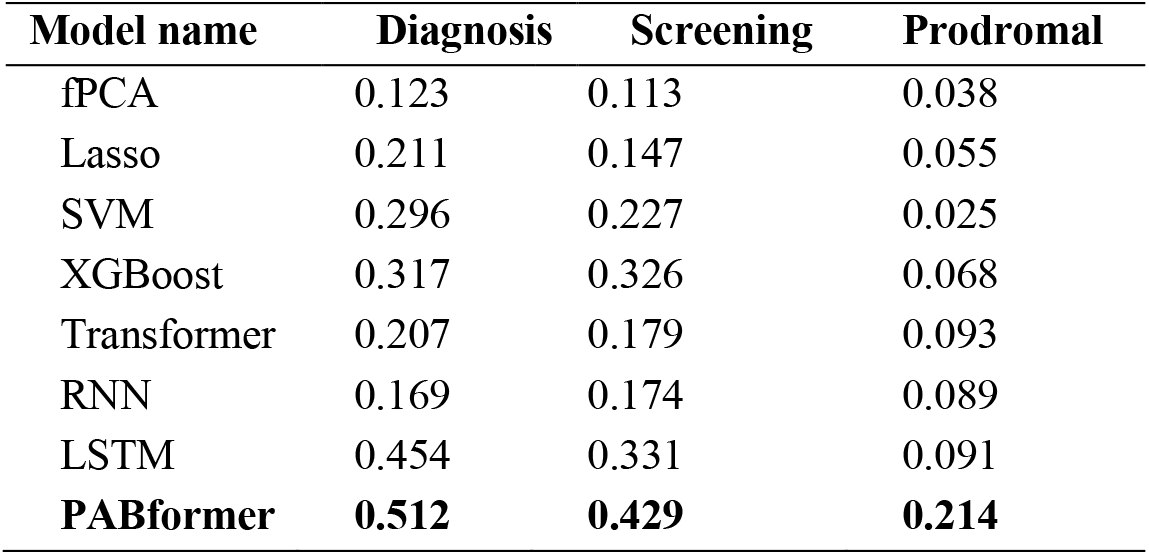
Performance comparison of models on arkinson’s disease classification tasks using PR AUC.

| Model name | Diagnosis | Screening | Prodromal |
| --- | --- | --- | --- |
| fPCA | 0.123 | 0.113 | 0.038 |
| Lasso | 0.211 | 0.147 | 0.055 |
| SVM | 0.296 | 0.227 | 0.025 |
| XGBoost | 0.317 | 0.326 | 0.068 |
| Transformer | 0.207 | 0.179 | 0.093 |
| RNN | 0.169 | 0.174 | 0.089 |
| LSTM | 0.454 | 0.331 | 0.091 |
| **PABformer** | **0.512** | **0.429** | **0.214** |

Traditional statistical and machine learning models perform substantially worse than PABformer, highlighting the limitations of handcrafted summary statistics for characterizing complex behavioral patterns. Deep learning models generally achieve stronger performance, indicating the importance of modeling temporal activity dynamics directly from accelerometer time series. Although the standard Transformer can model long-range temporal interactions, its performance remains inferior to PABformer, suggesting that preserving channel-specific behavioral information is beneficial for PD-related prediction tasks. Overall, these results demonstrate that PABformer provides a robust and generalizable framework for identifying both clinically manifest and prodromal Parkinson’s disease from wearable behavioral data.

### C. Behavioral pattern interpretation for PD diagnosis

We further investigated the interpretation of the PABformer model on the PD diagnosis task using the GradCAM method. GradCAM is a technique that provides visual explanations for deep neural networks by leveraging gradient information to highlight the most relevant regions in the image for a specific concept [46]. Higher GradCAM values indicate greater importance of the corresponding regions. Here, we adopted an aggregated GradCAM-based approach to visualize the channel importance by using the maximum importance scores across 100 randomly selected PD patients.

PABformer assigns high importance scores to spectral-based channels, followed by intensity-based channels, while statistics-based channels receive the lowest importance scores (Fig. 3a). Furthermore, we visualized the PA channels with high importance scores for individuals from different groups: prodromal patients, diagnosed patients, and healthy controls (Fig. 3b). Certain channels, such as SB count and MVPA count, are more important during the daytime and less important at night. The channel visualization in Fig. 3b also confirms that these channels show a large deviation of the disease group from the control group during daytime periods. Specifically, SB count increases in diagnosed and prodromal patients during the daytime, while MVPA count decreases.

**Fig. 3.**
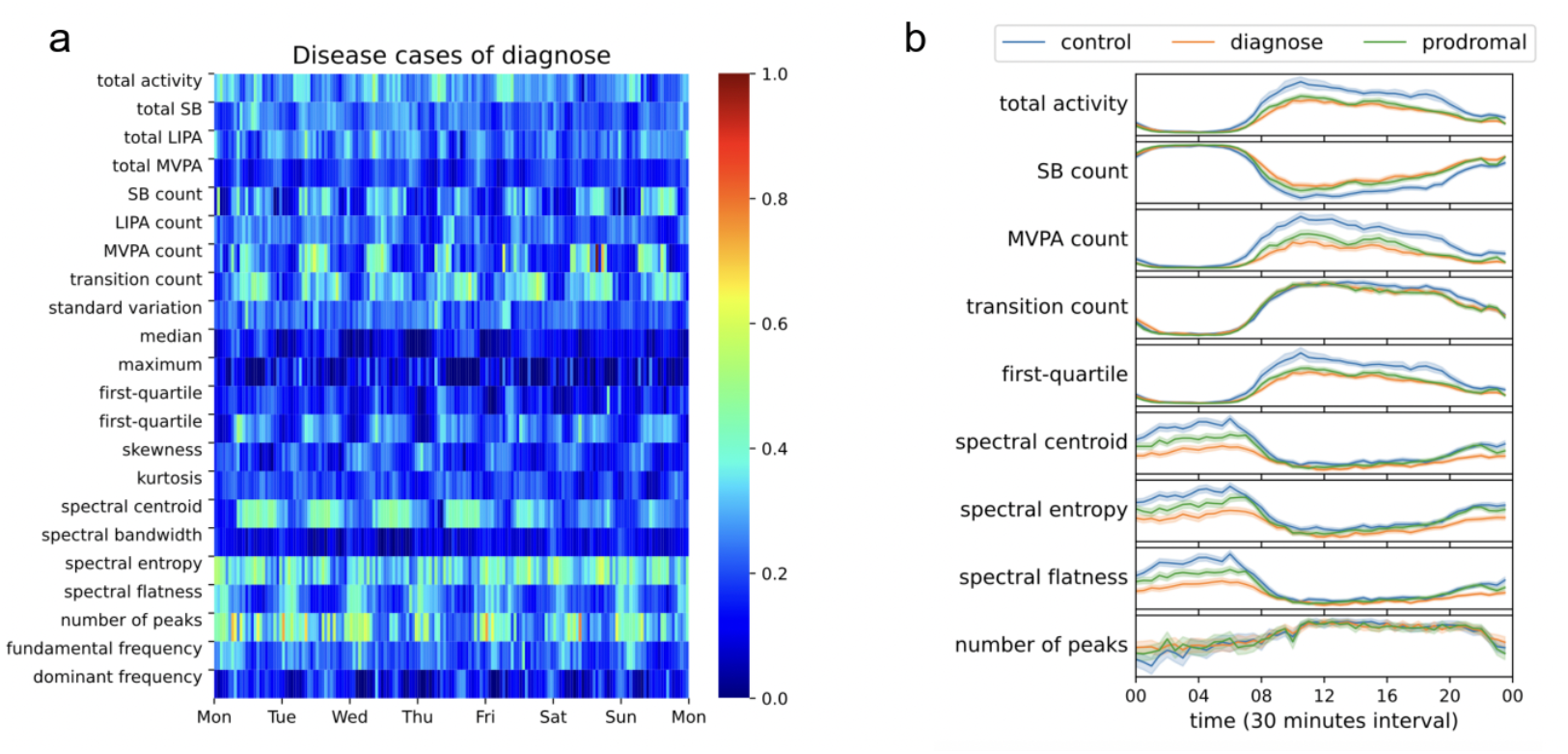
Interpretation of PABformer for PD diagnosis. (a) Visualization of channel importance learned by PABformer for PD diagnosis from GradCAM. (b) Visualization of important PA channels among the diagnostic, prodromal, and control groups. Shaded areas represent 95% confidence intervals of the estimated curves.

From the spectrum perspective, the spectral flatness channel demonstrated greater importance at night than during the daytime (Fig. 3a). Spectral flatness quantifies the degree to which a signal resembles white noise, the spectral centroid represents the “center of mass” of the frequency spectrum, and spectral entropy measures the randomness or complexity of the PA signal. Among patients with PD, the night values of spectral centroid, entropy, and flatness were much lower than those in the control group (Fig 3b). These lower values suggest reduced movement complexity and diminished variability in nocturnal activity patterns among individuals with PD. Such alterations may be related to impaired bed mobility (IBM), a common nocturnal symptom of PD characterized by difficulty intentionally repositioning the body during sleep [54]. Importantly, prodromal individuals already exhibited behavioral patterns that deviated from healthy controls and partially resembled those observed in diagnosed PD patients, providing a potential explanation for the strong performance of PABformer in prodromal PD prediction. These results indicate that PABformer captures clinically meaningful behavioral patterns associated with PD [54].

### D. Incident PD prediction

We evaluated the predictive performance of PABformer on incident PD using the Cox proportional hazards (Cox PH) survival model. The model was fine-tuned to predict time-to-event outcomes, where the objective was to estimate the risk of future PD incidence based on accelerometer-derived behavioral representations. Model performance was assessed using the concordance index (C-index) and inverse probability of censoring weighted concordance index (IPCW C-index) to evaluate risk ranking ability, as well as the 5-year time-dependent ROC AUC to assess fixed-horizon prediction performance.

As shown in Fig. 4a, the combined model (PABformer + covariates) achieved the best performance across all metrics, with a C-index of 0.866, an IPCW C-index of 0.867, and a 5-year ROC AUC of 0.870, substantially outperforming the covariate-only baseline (0.746, 0.742, and 0.733, respectively). These results demonstrate that wearable-derived behavioral representations contain potentially informative preclinical signals associated with future PD onset. Notably, the large improvement from the covariate-only baseline to the PABformer-only model indicates that accelerometer time-series data provide predictive information beyond traditional clinical risk factors. Furthermore, the additional performance gain achieved by integrating covariates with PABformer suggests that behavioral representations and conventional demographic or lifestyle variables provide complementary information for PD risk prediction. The consistently high IPCW-adjusted concordance values also indicate that the model remains robust under censored longitudinal follow-up conditions. In addition, the strong 5-year ROC AUC performance demonstrates that the learned representations retain predictive value several years prior to formal clinical diagnosis, highlighting the potential utility of PABformer for early PD risk stratification and long-term disease surveillance.

**Fig. 4.**
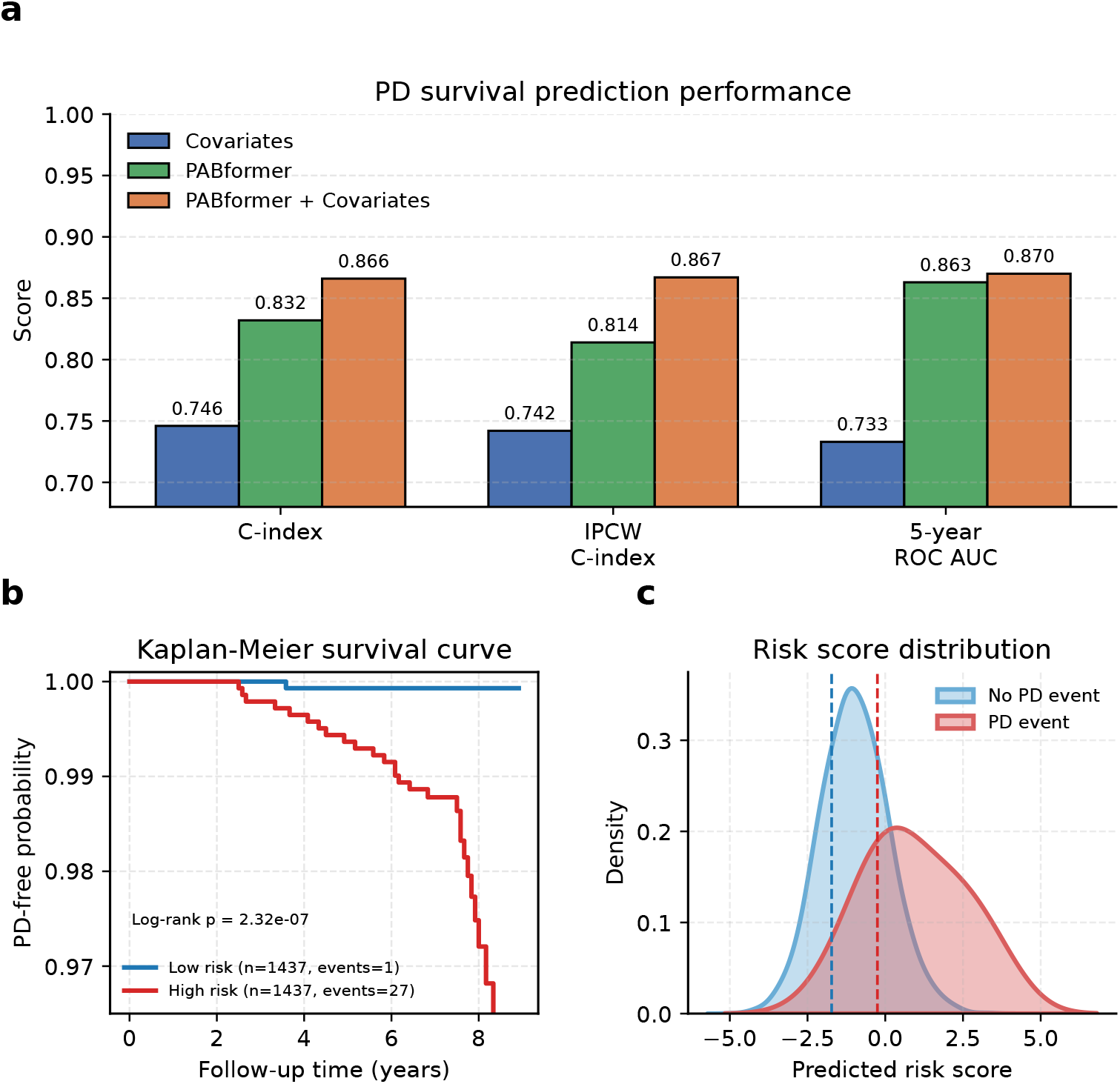
Incident PD prediction using PABformer. (a) Comparison of survival prediction performance between covariate-only, PABformer-only, and combined models. (b) Kaplan–Meier curves comparing PD-free survival between high-risk (top 25%) and low-risk (bottom 25%) groups stratified by predicted risk scores. (c) Distribution of predicted risk scores for individuals with and without future PD events. Dashed lines denote the 25th and 75th percentile cutoffs of the predicted risk scores in the test set, which were used to define the low-risk and high-risk groups for Kaplan–Meier analysis.

To evaluate the model’s clinical utility, individuals were stratified into high- and low-risk groups based on predicted risk scores. The Kaplan–Meier survival curves (Fig. 4b) show a clear separation between the two groups. The high-risk group exhibited a substantially faster decline in PD-free survival probability compared with the low-risk group, indicating markedly elevated future PD incidence among individuals assigned higher predicted risk scores. The separation between the two survival curves became increasingly apparent as follow-up progressed, suggesting that the predicted risk scores capture behavioral patterns associated with future disease development. The difference between the two groups was statistically significant (log-rank *p* = 2.32 × 10^*−7*^), confirming that the predicted risk scores are highly informative of future PD incidence.

We further examined the distribution of predicted risk scores (Fig. 4c). Individuals who developed PD showed a clear shift toward higher predicted risk scores compared with non-event individuals, with relatively limited overlap between the two distributions. In contrast, non-PD individuals were concentrated in the lower-risk region. This separation demonstrates that PABformer provides meaningful risk stratification at the individual level and is capable of distinguishing subtle behavioral differences prior to clinical diagnosis.

Overall, these results demonstrate that PABformer not only achieves strong predictive performance in terms of concordance metrics but also provides clinically interpretable risk stratification. The combination of high C-index, significant Kaplan–Meier separation, and well-separated risk distributions highlights the effectiveness of PABformer in capturing predictive patterns from accelerometer time-series data for incident PD prediction.

## V. Discussion

In this study, we proposed PABformer, a multi-channel Transformer framework pretrained on large-scale accelerometer data from the UK Biobank to learn comprehensive PA behavior representations. By leveraging the Transformer architecture’s ability to capture long-range temporal dependencies and introducing a channel-separation strategy to disentangle heterogeneous PA signals, PABformer effectively models complex and dynamic behavioral patterns from week-long accelerometer recordings. Using data from 81,463 participants for pretraining, we fine-tuned PABformer for downstream tasks including demographic attribute inference, PD diagnosis, and incident PD prediction. Compared with traditional statistical and machine learning, recurrent deep learning models, and a standard Transformer without channel separation, PABformer consistently achieved superior performance in both PD classification and survival prediction tasks, demonstrating its ability to learn more informative representations than conventional PA summary statistics and existing computational approaches. Furthermore, the integration of Grad-CAM improved model interpretability by identifying salient channel contributions, enhancing transparency and providing potentially clinically meaningful insights into PD behavioral patterns. Compared with previous foundation-model approaches for accelerometer data, such as Pretrained Actigraphy Transformer (PAT) [38], which operates on single-channel data, PABformer explicitly models the multi-dimensional structure of accelerometer signals through channel separation. Combined with large-scale pretraining, this design enhances the model’s ability to capture heterogeneous activity patterns, improving both robustness and generalizability.

We provided an open-source implementation of PABformer on GitHub (https://github.com/mengwang-lab/PABformer), comprising a pretraining module and a fine-tuning module that support a variety of downstream tasks. For users with access to large-scale accelerometer datasets from specific populations, the pretraining module can be used to learn PA behavior representations. For users without large datasets but with a focus on a downstream task, the fine-tuning module enables transfer learning from a pretrained model trained on the UK Biobank. This setup highlights the advantage of foundation models in low-sample regimes, where leveraging a pretrained model typically yields better performance than training models from scratch. The fine-tuning module is flexible, allowing users to incorporate additional vectorized covariates—such as blood biochemistry biomarkers—via simple concatenation with the PA behavior representations. For downstream analysis, we support regression, classification, and survival prediction tasks (via Cox PH models), which can be easily specified through task selection.

The PA behavior representations learned by PABformer can be seamlessly integrated into multimodal modeling frameworks for comprehensive health outcome prediction. These representations, capturing high-resolution temporal dynamics of physical activity, can serve as one modality alongside other data types such as genomics, imaging, electronic health records, and biochemical markers. For downstream multimodal tasks, the PA behavior embeddings can be concatenated with vectorized features from other modalities or integrated using more advanced multi-modal fusion techniques [55]. Given the modular design of PABformer, it can flexibly interface with existing multimodal pipelines and serve as a plug-and-play component for longitudinal disease risk modeling and precision health applications.

Integrating additional cohorts into the training of PABformer presents a promising direction to improve model generalizability and mitigate the demographic and geographic biases inherent in single-cohort training. By incorporating accelerometer data from diverse populations, PABformer could learn more robust and transferable PA behavior representations across age groups, ancestries, and lifestyle contexts. However, several challenges must be addressed to enable successful multi-cohort integration. First, heterogeneity in accelerometer device types, wear protocols (e.g., wrist vs. hip placement), sampling frequencies, and data preprocessing pipelines can introduce batch effects that confound representation learning. Second, harmonizing outcome definitions and covariate availability across studies remains nontrivial, particularly for incident disease phenotyping. Third, privacy and data governance constraints may limit centralized model training, making federated learning approaches [56] an appealing alternative. Future work could integrate PABformer into a privacy-preserving federated learning framework combined with data harmonization strategies to mitigate device and protocol differences, thereby enhancing model robustness and generalizability.

## VI. Limitations

This study has several limitations. First, the accelerometer data were derived exclusively from the UK Biobank, whose participants are predominantly middle-aged to older adults of White British ancestry. This demographic composition may limit the generalizability of the proposed framework to more diverse populations and healthcare settings. Future work will focus on integrating data from multiple cohorts through privacy-preserving and federated learning strategies to improve model robustness and population-level applicability. Second, this study primarily focused on PD diagnosis and prediction, but the proposed framework is generalizable to other health outcomes and chronic diseases. Incorporating additional biomarkers, clinical variables, or multimodal data may further enhance predictive performance and clinical utility. Finally, PABformer was developed using 5-second epoch accelerometer data, following the standard preprocessing pipeline of the UK Biobank. Future studies are needed to evaluate the impact of alternative preprocessing strategies, epoch lengths, and device-specific formats on model performance, reproducibility, and cross-cohort generalizability.

## VII. Conclusion

In conclusion, we developed PABformer, a scalable and generalizable foundation model for accelerometer data that effectively learns rich physical activity behavior representations from long-term recordings. By capturing complex temporal and heterogeneous behavioral patterns, PABformer enhances the utility of accelerometer measurements for Parkinson’s disease diagnosis and prediction, outperforming conventional summary-statistic approaches, traditional statistical and machine learning methods, recurrent neural networks and Transformer-based baselines. These findings highlight the potential of foundation models for wearable sensor data to advance early disease detection, improve risk prediction, and support future precision health applications.

## Data Availability

All accelerometer, phenotype, and health outcome data used in this study were obtained from the UK Biobank. Access to these data is available through the UK Biobank Research Analysis Platform (https://www.ukbiobank.ac.uk/) upon application and approval.

